# An LLM-assisted Pipeline for Identifying ClinicalTrials.gov Registry Changes and Their Association with Primary Endpoint Positivity

**DOI:** 10.64898/2026.11.17.26368319

**Authors:** Ayurzana Purevdorj, Fabio Dennstädt, Daniel R. Zwahlen, Paul Windisch

**Author notes:** Corresponding author: Ayurzana Purevdorj, MD Department of Radiation Oncology Kantonsspital Winterthur Brauerstrasse 15, Haus R, 8400 Winterthur.

## Abstract

Registration of randomized clinical trials (RCTs) in a public registry such as ClinicalTrials.gov should minimize the risk of reporting bias. However, records may be modified during a trial and the resulting discrepancies are not always apparent without a detailed examination of all version histories, a process that is cumbersome when performed manually. We developed an automated workflow to investigate whether specific changes to RCT registry entries are associated with primary endpoint positivity. Our pipeline extracts RCT records from ClinicalTrials.gov and systematically detects changes between successive versions using a combination of deterministic textual analysis and large language model (LLM)-assisted classification. We could show a positive association between trial positivity and changes to primary outcomes (OR 1.91; 95% CI 1.48-2.47) as well as to arms and interventions (OR 2.10; 95% CI 1.62-2.73). Associations were weaker when changes occurred exclusively before the primary completion date, pointing to the importance of timing in evaluating potentially consequential modifications to RCT design. Our automated pipeline can enable analysis of trial registration records at scale, facilitating the work of reviewers and meta-scientists to promote research integrity.

## Introduction

Prospective registration of clinical trials in a publicly accessible database such as ClinicalTrials.gov^1^ is considered an essential part of good research practice, enhancing transparency and accountability in the reporting of clinical trials. It is ethically required by the Declaration of Helsinki for every study involving human subjects^2^, and is legally mandated for certain categories of trials in some jurisdictions^3^.

Beginning in 2005, the International Committee of Medical Journal Editors (ICMJE) required investigators to register a clinical trial before patient enrollment as a precondition for publication in its journals^4^, resulting in a marked increase in the number of registered trials.

Prospective registration helps minimize the risk of publication bias and outcome reporting bias, as long as outcomes and other trial characteristics are reported completely and accurately in a register. It can also facilitate the work of editors and peer reviewers in verifying that there are no discrepancies between study registration records and published results, and aid the evaluation of biomedical research conducted by systematic reviewers and meta-researchers. Finally, it fulfills an essential function for the public by providing information about new and ongoing trials.

Nevertheless, methodologically questionable practices still occur^5^, and in-depth research of study registration records can sometimes help expose discrepancies which are not apparent from published results. Still, identifying changes in registry data -especially for a large number of trials - is cumbersome given that some trials undergo a substantial number of changes in their records, resulting in numerous registry versions. Both manual and automated approaches to registry history analysis have been employed in the literature.

The largest clinical trial registry is ClinicalTrials.gov, launched in February 2000. Several studies have examined inconsistencies among records in this registry, mainly focusing on outcome changes. Most investigators have assessed discrepancies between primary outcomes reported in registry entries and those reported in subsequent publications^6–10^. A systematic review of meta-research studies found that such discrepancies occurred in a median of 31% of trials^11^. Some authors have also analyzed within-registry changes of primary outcomes over the course of a trial’s history^12–14^, though this approach was often limited to a pairwise comparison between two versions for each trial (for example between the earliest submitted and the latest available entry). Only in recent years have tools been developed to automatically retrieve complete registry histories from ClinicalTrials.gov and conduct a longitudinal analysis of modifications across an entire trail of study records^15^. Holst et al. used one such approach and assessed the frequency of primary outcome changes in a cohort of German trials^10^, observing that a substantial proportion of changes would not be apparent if limiting the search to a comparison between the earliest and the current registry entry.

In addition, some investigators have analyzed measures of possible outcome reporting bias and found a tendency of articles with discrepancies between registered and published outcomes to report statistically significant results^8^. One study evaluated the association between primary outcome changes and reported intervention effect size, finding that trials with such changes were more likely to report larger intervention effects than trials without^6^.

To the best of our knowledge, no prior investigation has systematically evaluated longitudinal changes in multiple registry fields beyond primary outcomes across the complete registry history of a cohort of trials.

Our objective in this study was twofold: (a) to develop an automated workflow capable of retrieving complete registry histories and identifying changes across successive record versions, and (b) to explore whether these registry changes are associated with the likelihood of a positive primary endpoint.

## Methods

### Data selection

The raw dataset consists of all articles containing an abstract drawn from four major medical journals (the New England Journal of Medicine, JAMA, the British Medical Journal and the Lancet) that were published between January 1, 2020 and December 31, 2025 (n = 7552). The abstracts were retrieved as a plain text file from MEDLINE through the PubMed interface.

### LLM-based abstract classification

The GPT-5.6 model (OpenAI, San Francisco, CA, USA), Sol version, was used via its vendor API in a local pipeline. The reasoning level was set at medium, other parameters were left under their default vendor configuration as verified on August 4, 2026 (temperature =1.0, top_p = 0.98).

In the first stage, publication abstracts were screened by the model to identify randomized controlled trial (RCT) results reports. Primary results publications as well as secondary-endpoint, follow-up, and post-hoc analysis publications were considered eligible as long as they preserve the comparison between randomized groups. Protocol publications, editorials, reviews, nonrandomized studies as well as meta-analyses and RCT-derived analyses that pool RCTs or analyze only one arm were excluded.

Publications were assigned a label through a structured output schema: “primary rct report”, “not primary rct report”, “unclear”. The abstracts labeled as “primary rct reports” were parsed using regular expressions to extract the corresponding National Clinical Trial (NCT) number.

A downstream workflow also using GPT 5.6 Sol at the same settings classified the publications previously identified as RCT reports according to their primary endpoint. The abstracts were labeled as “positive”, “negative” or “unclear” according to whether the primary endpoint met the trial’s stated success criterion in the intended direction. For multiple primary endpoints an explicitly stated overall success rule was needed to assign a positive label. Mixed results without such a rule were labeled as “unclear”. The exact prompts that were used are provided in Supplementary Table 1 and 2.

Even though it has been previously demonstrated that LLMs are very capable at performing such a task^16^, we conducted a verification of the LLM-based classification on a randomly drawn sample of 50 abstracts. Two physicians (A.P. and P.W.) independently annotated the abstracts; annotation results were compared between the human annotators and between each human annotator and the LLM using Cohen’s kappa (κ).

### Retrieval and Comparison of ClinicalTrials.gov Study Record Versions

Trials identified as RCTs and containing an NCT number were processed through a separate pipeline developed to retrieve and compare historical versions of their ClinicalTrials.gov registry records.

This deterministic Python pipeline queried ClinicalTrials.gov application programming interfaces (APIs) to validate NCT identifiers, retrieve current study records along with every available public registry version, obtain metadata and deduplicate publications referring to the same NCT trial.

From these historical versions we extracted specific registry fields contained in the sections “Study Design”, “Arms and Interventions”, and “Outcome Measures”, as well as landmark dates such as the study-start date. Within those fields we distinguished between scalar (i.e. data types which only contain a single value) and nested items. Scalar items (such as study type, allocation, masking etc.) were captured by taking a snapshot of that field at a specific version in time. Nested items are domains with repeated, hierarchical, or text-rich content (such as outcomes, study arms, etc.), where representation in single scalar columns would lead to information loss. These were extracted into separate version-specific tables, and a conservative text normalization process was performed, whereby whitespaces were trimmed or collapsed, and letter case was standardized.

All consecutive study record versions within each NCT trial were then compared pairwise. Each change date was also compared to trial-specific time anchors such as study start and primary completion date and a corresponding indicator was generated.

When comparing scalar or nested fields between versions, a classification label with the possible values “added”, “removed”, “modified” or “reordered” was generated. For scalar fields “added” meant that a previously empty field became populated, “removed” meant the reverse, and “modified” meant a change in its value. For nested fields a textual matching procedure was first performed; a changed attribute of a matched item resulted in the “modified” label, whereas an unmatched item in the earlier or later version was recorded as “removed” or “added” respectively. Changes in item order alone were recorded as “reordered”. In addition, outcomes could also be labeled as “reclassified” when a change in priority occurred. As a result of item matching, registered changes were recorded in nested changes csv tables, where one single matched item could produce multiple rows of changes. For details about the matching procedure, see Supplementary Table 3.

Selected changes and observations were then flagged using deterministic rules. We defined eleven flag categories (see Table 1 and 2).

**Table 1.** Main deterministic review flags. Eleven main flag categories drawn from the ClinicalTrials.gov Study Plan section and from Study Record Dates were chosen.

| **Flag code** | **Deterministic trigger** |
|---|---|
| **STUDY TYPE CHANGED** | Study type is added, removed, or changed between consecutive public versions. |
| **ALLOCATION CHANGED** | Allocation is added, removed, or changed. |
| **INTERVENTION MODEL CHANGED** | Intervention model is added, removed, or changed. |
| **PRIMARY PURPOSE CHANGED** | Primary purpose is added, removed, or changed. |
| **MASKING CHANGED** | Masking category is added, removed, or changed. |
| **NUMBER OF ARMS CHANGED** | Number of arms is added, removed, or changed. |
| **PLANNED ENROLLMENT CHANGED** | Enrollment count changes with both versions classifying enrollment as ESTIMATED. Estimated-to-actual and actual-to-actual transitions are excluded. |
| **PRIMARY OUTCOME CHANGED** | A primary outcome is added, removed, modified, or reclassified. Description-only changes are excluded, as is pure reordering. |
| **ARMS INTERVENTIONS CHANGED** | An arm or intervention item is added, removed, or modified. Description-only and pure ordering changes are excluded. |
| **FIRST POST AFTER STUDY START** | The earliest public posting date is later than the earliest reported study-start date. |
| **START DATE CROSSED FIRST SUBMISSION BOUNDARY** | Between any consecutive versions, the reported start date moves from before first submission to on/after first submission, or the reverse. |

**Table 2.**
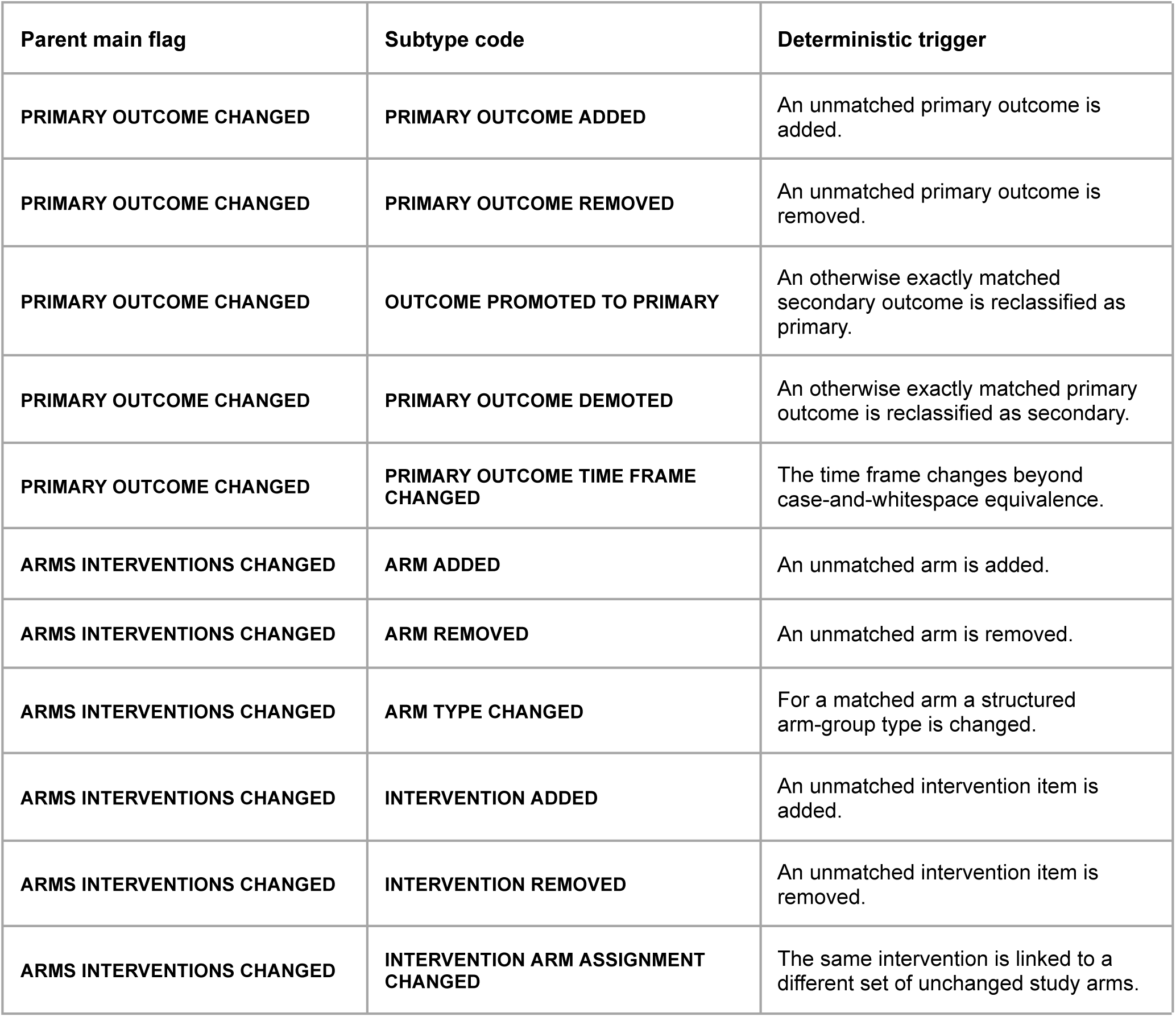
Nested change subtypes.

| **Parent main flag** | **Subtype code** | **Deterministic trigger** |
|---|---|---|
| **PRIMARY OUTCOME CHANGED** | **PRIMARY OUTCOME ADDED** | An unmatched primary outcome is added. |
| **PRIMARY OUTCOME CHANGED** | **PRIMARY OUTCOME REMOVED** | An unmatched primary outcome is removed. |
| **PRIMARY OUTCOME CHANGED** | **OUTCOME PROMOTED TO PRIMARY** | An otherwise exactly matched secondary outcome is reclassified as primary. |
| **PRIMARY OUTCOME CHANGED** | **PRIMARY OUTCOME DEMOTED** | An otherwise exactly matched primary outcome is reclassified as secondary. |
| **PRIMARY OUTCOME CHANGED** | **PRIMARY OUTCOME TIME FRAME CHANGED** | The time frame changes beyond case-and-whitespace equivalence. |
| **ARMS INTERVENTIONS CHANGED** | **ARM ADDED** | An unmatched arm is added. |
| **ARMS INTERVENTIONS CHANGED** | **ARM REMOVED** | An unmatched arm is removed. |
| **ARMS INTERVENTIONS CHANGED** | **ARM TYPE CHANGED** | For a matched arm a structured arm-group type is changed. |
| **ARMS INTERVENTIONS CHANGED** | **INTERVENTION ADDED** | An unmatched intervention item is added. |
| **ARMS INTERVENTIONS CHANGED** | **INTERVENTION REMOVED** | An unmatched intervention item is removed. |
| **ARMS INTERVENTIONS CHANGED** | **INTERVENTION ARM ASSIGNMENT CHANGED** | The same intervention is linked to a different set of unchanged study arms. |

Flags are generated by all types of changes (with the exception of mere reordering) in the following scalar fields: study type, allocation, intervention model, primary purpose, masking, and number of arms.

An enrollment-count change is flagged only if a planned enrollment change occurs (i.e. a change from an estimated enrollment count to a new estimated count).

Changes to the nested fields “primary outcomes” and “arms and interventions” also generate a flag; in the case of primary outcomes only changes to the following components (data elements) trigger a flag: “Outcome measure/type” and “Outcome time frame”, whereas textual changes to “Outcome description” are ignored. In the case of arms and interventions, textual changes to “Arm description” or “Intervention description” are likewise ignored. This was chosen in order to increase specificity for substantive changes and reduce the number of flags triggered by minor modifications.

For nested changes additional flag subtypes were assigned, according to whether an item was added, removed, modified or an outcome priority change occurred (secondary outcome promoted to primary or vice versa).

A specific flag is also assigned in two cases relating to study dates: if the earliest public posting of a clinical trial occurs after the earliest reported study-start date, and if, comparing two consecutive versions, the reported start date of the study moves from before first submission to on/after first submission, or vice versa (change from retrospective to prospective trial registration, or vice versa).

The pipeline finally generates comparison outputs in the form of csv tables for each flagged change.

### Timing analysis relative to primary completion date

To assess the impact of the timing of changes on our association analysis we divided the trials in groups according to whether a change took place exclusively before or at primary completion date (defined as the “early only” group) or exclusively after this date (“late only” group); we also considered trials having changes both before and after primary completion, as well as different combinations of these groups (groups “any before” and “any after”).

### LLM-based outcome change assessment

To improve discrimination capacity for identifying substantive changes of primary outcomes, an automated LLM-assisted semantic assessment module was developed in parallel to the deterministic pipeline.

This layer employed the same model described above (OpenAI GPT-5.6 Sol via API at reasoning level medium). A single API call per trial was made, using one prompt containing the instructions as well as all historical versions of primary outcomes of that trial extracted from ClinicalTrials.gov. The prompt instructs the LLM to review every consecutive pair of registry entry versions as well as the first with the last one and to assess whether a meaningful change to the primary outcome occurred. Meaningful changes to the primary outcome were defined as those changes which may substantially affect the probability of obtaining a positive result. More specifically, the following were considered meaningful changes: addition or removal of an outcome; modifications to outcome definition, assessment time frame, measurement method, or threshold/cutoff; primary outcome demotion to secondary or secondary outcome promotion; outcome replacement (complete change in outcome characteristics not accompanied by addition or removal of a second outcome). Semantically equivalent rewording as well as mere spelling, punctuation, and formatting changes were thus excluded. Furthermore, adding or removing clarifications which do not substantially affect the above-mentioned outcome components was not considered a meaningful change.

In order to identify changes to outcome priority a deterministic upstream layer was employed. This selects a candidate secondary outcome for matching according to text similarity rules (word count cosine similarity and Jaccard distance) and adds it to the prompt; the LLM establishes then whether an outcome promotion or demotion has occurred.

The LLM was instructed to deliver one structured output per trial containing a Boolean label for the presence of meaningful changes (“True” or “False”), the exact version transition containing that change, and a “change type” label corresponding to one of the above-mentioned categories.

### Statistical Analysis

We described categorical variables with the use of frequencies and percentages. Associations between the presence of flagged changes at the trial level and primary endpoint positivity were estimated by calculating crude odds ratios (OR) with an exact 95% confidence interval (CI) and a two-sided Fisher exact test. We also estimated the association of changes with endpoint positivity by fitting a Firth penalized adjusted logistic regression model, with endpoint positivity as the dependent variable. Adjustment was made for three prespecified covariates: calendar year of first submission to ClinicalTrials.gov per five-year increase, trial phase (grouped as early phase = phase 1, phase 1/2, phase 2; late phase = phase 2/3, phase 3; post-marketing = phase 4; not applicable; missing), and initial planned enrollment, per doubling on a base-2 logarithmic scale. We adjusted for first-submission year to account for historical trends in registration practices and because older records may have had a longer opportunity to accumulate updates. Trial phase was included to account for differences in study objective and trial design. Initial planned enrollment as a measure of intended trial size was included because of possible size-related differences in statistical precision and trial complexity. Adjusted ORs were reported with 95% profile penalized-likelihood CIs and penalized likelihood-ratio p values. These were purely exploratory analyses and intended to quantify associations rather than infer causal effects. Statistical analyses were conducted using the software R version 4.5.3 (R Foundation for Statistical Computing, Vienna, Austria).

## Results

### RCT classification and NCT number extraction

7552 publication abstracts were extracted via PubMed according to our selection criteria. (See Fig. 1). Our LLM-based pipeline for RCT classification identified 2178 paper abstracts reporting results of RCTs. 5352 publications were classified as not reporting RCTs, and 22 were labeled as “unclear”, meaning that definitive attribution was not possible.

**Fig. 1.**
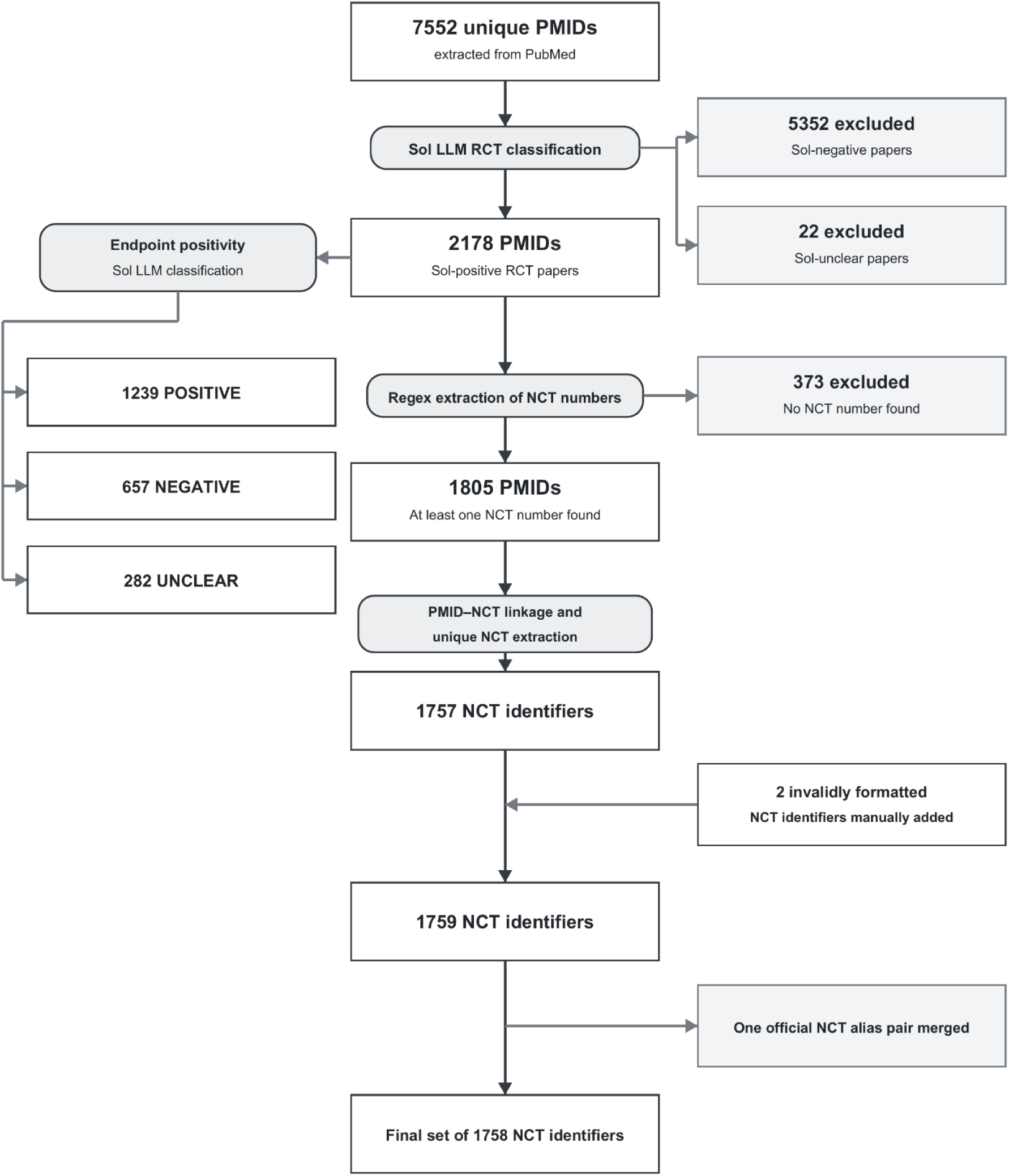
Flowchart of RCT classification and NCT number extraction.

The second stage of the LLM-classification analyzed the abstracts of the 2178 RCT-positive articles and determined 1239 publications to have met a positive primary endpoint, 657 a negative one. For 282 publications the primary endpoint success could not be definitively established from the abstract alone.

The concordance for RCT classification between GPT-5.6 and the human annotators AP and PW, as measured on the random 50-article sample, was 100% for both annotators (Cohen’s κ = 1.0). The concordance for primary endpoint positivity between the LLM and annotator AP was 100% (Cohen’s κ = 1.0), and between the LLM and annotator PW was 88.24% (Cohen’s κ = 0.76).

Through regular expression search 1805 abstracts were found to have at least one NCT identifier. 371 abstracts contained either an alternative clinical trial registry number (334 cases), or no registry number at all (37 cases); moreover, 2 abstracts contained an invalidly formatted NCT registration number. Subsequently the linkage between publications and NCT numbers was analyzed to resolve cases in which a single publication contained multiple NCT numbers, or a single NCT trial was represented by multiple publications.

In this manner 1757 unique NCT identifiers were extracted. The two invalidly formatted NCT numbers were manually added to the set. Finally, one NCT number was removed as it was part of an NCT alias pair referring to the same trial. This resulted in a final set of 1758 NCT trials, which formed the basis for our subsequent analysis.

### Descriptive analysis

Our deterministic pipeline established the presence of 18 024 individual flagged changes within all historical registry versions of the NCT trials (Figure 2 a-b). Among these, 16 054 changes were in nested fields (6978 primary outcome changes and 9076 arms and interventions changes), whereas 1970 changes were non-nested (including 1286 scalar field changes and 684 changes of the type “First public posting after study start”).

**Fig. 2.**
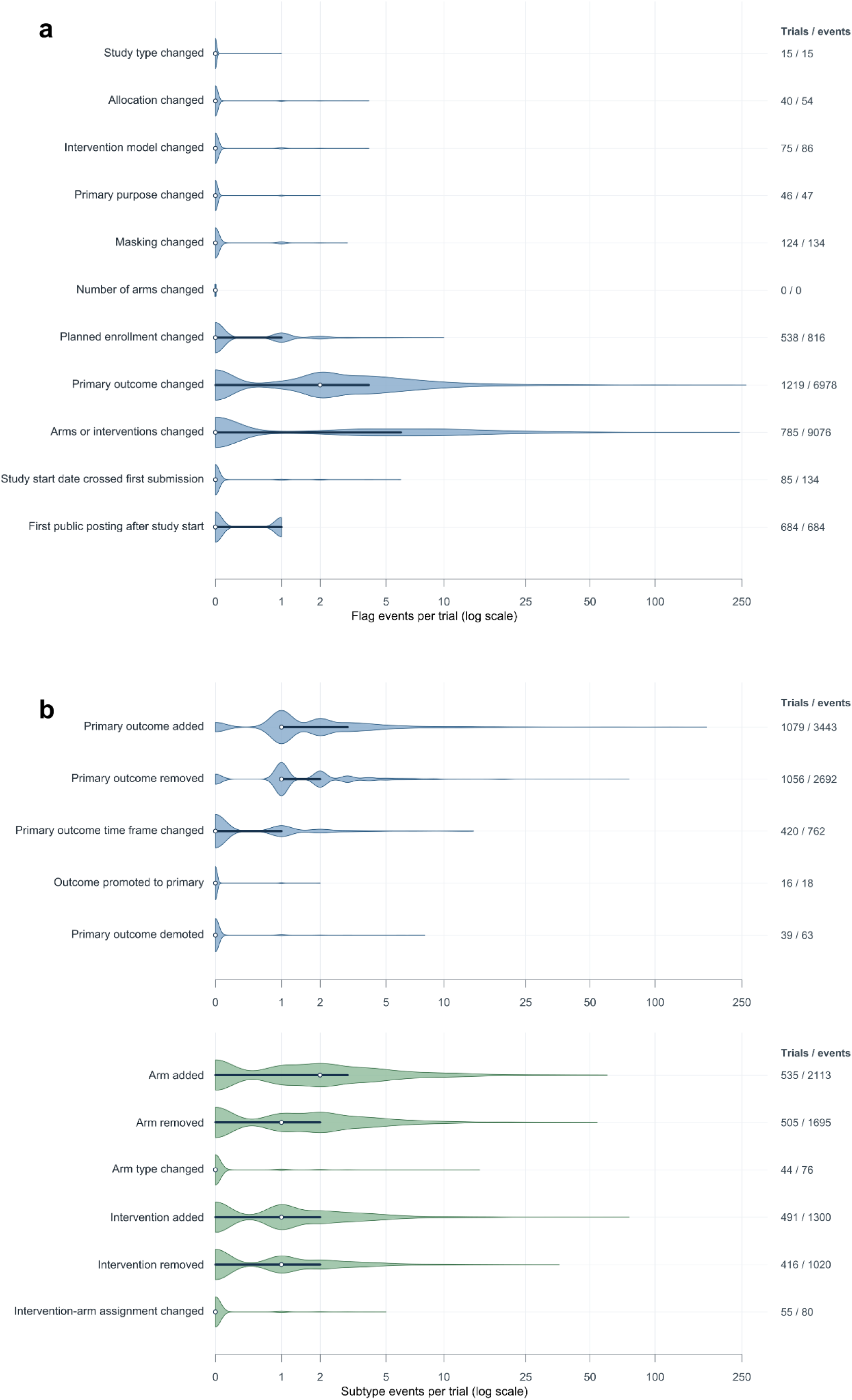
Distribution of the number of changes per trial. (A) all flagged changes, stratified by flag category; (B) nested-field changes, stratified by flag category subtype point. White dot: median; dark line: interquartile range.

In the scalar field categories almost all trials had either zero or one change. On the other hand, the nested field categories showed a markedly right-skewed distribution, with the top outlier trials having up to 262 primary outcome changes and 244 arms and interventions changes.

Flag categories showed a variable prevalence at the trial level (Fig. 3 a-b), ranging from 0 (there were no trials having a change in the number of arms) to 69.3% (prevalence of primary outcome changes across all historical registry versions).

**Fig. 3.**
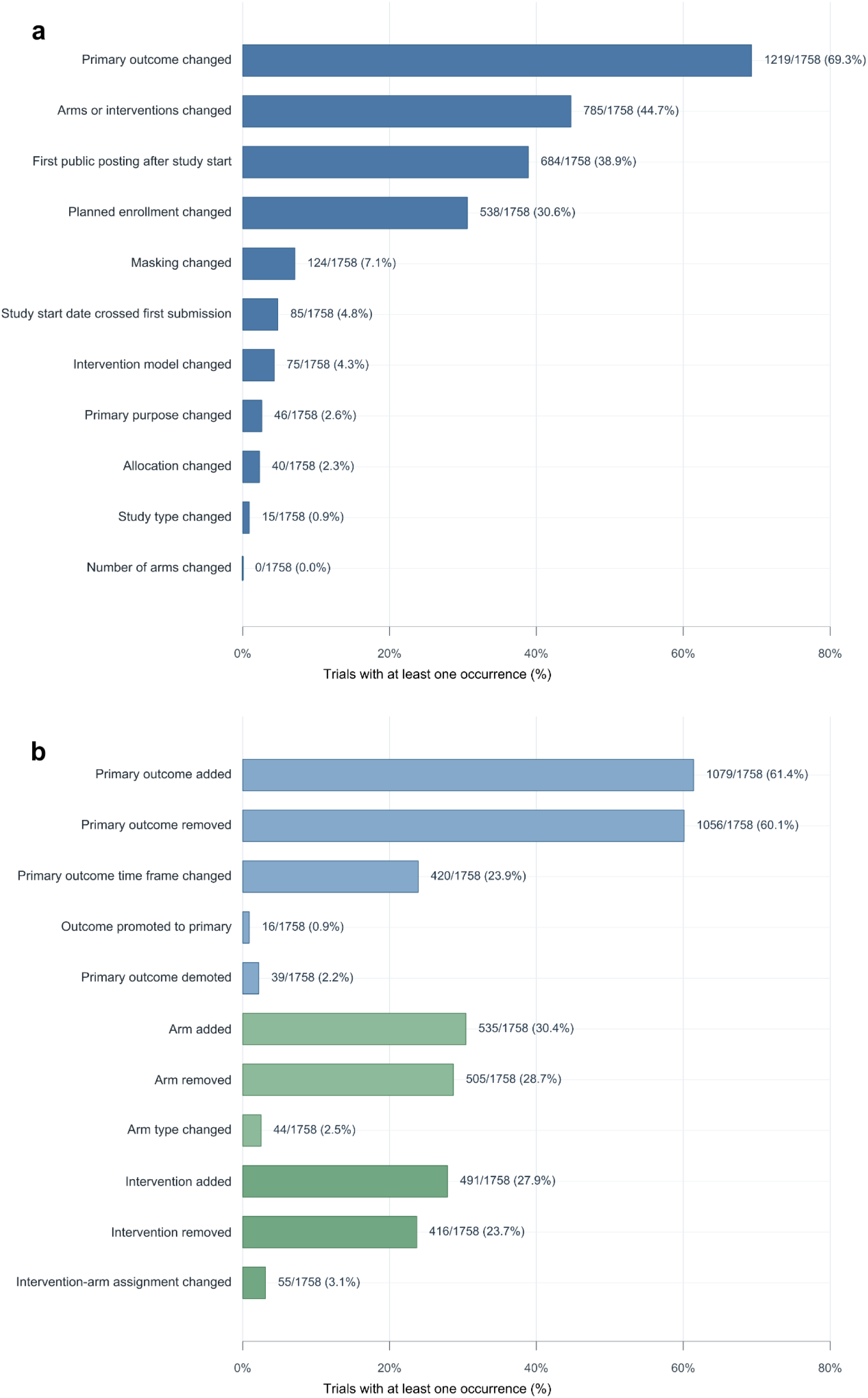
Trial-level prevalence of flag categories (3a) and nested field change subtypes (3b).

Our analysis included two registration-timing flag categories. “First public posting after study start”, which is equivalent to the general definition of retrospective registration, included 684 trials, of which 390 were definitively first registered after study start, and 294 had a start date reported with month precision, potentially overlapping with the submission date.

“Start date crossed first submission boundary” signals that a trial underwent a transition from a retrospective to a prospective registration, or vice versa, or both. There were 85 flagged trials in total, with 46 trials having retrospective-to-prospective transitions only, 5 having prospective-to-retrospective transitions only, and 34 having both.

### Association with endpoint positivity

We aimed to determine the association of flagged trial record changes with primary-endpoint positivity. Since the outcome of our analysis was binary (primary endpoint either positive or negative), we had to restrict our set of NCT trials by omitting ambiguous cases. From the original set of 1758 trials with a unique NCT identifier we excluded 193 trials whose associated abstracts were classified as “unclear” regarding primary endpoint success by the LLM. In addition, we excluded 12 trials associated with multiple publications which reported both positive and negative primary endpoints. We further excluded 110 trials because they were linked to abstracts citing multiple NCT trials, making it unclear whether the label “positive endpoint” could be applied to all corresponding trials. Finally, since we conducted an adjusted regression analysis accounting for planned enrollment count as a covariate, we also excluded 43 trials lacking a planned enrollment in their first available registry entry. This led to a final set of 1400 trials, of which 967 (69.1%) had a positive primary endpoint and 433 (30.9%) a negative one.

A particularly strong association was found for changes in primary outcome as well as in arms and interventions (Fig. 4), showing an unadjusted OR of 1.94 (95% CI 1.52-2.48) and 2.15 (95% CI 1.68-2.77) respectively. This association was confirmed in the adjusted logistic regression analysis, with a corresponding OR of 1.91 (95% CI 1.48-2.47) for primary outcome changes and 2.10 (95% CI 1.62-2.73) for arms and interventions changes. We were also interested in isolating the association between endpoint positivity and the transition from a retrospective to a prospective registration specifically, so we included it as a separate flag (Start date: retrospective to prospective only) in our analysis. Both the flag “Study start date crossed first submission” and its subset of retrospective to prospective only transitions showed a tendency towards association with endpoint positivity (unadjusted OR 1.46 [0.79, 2.83] and 1.88 [0.80, 5.13] respectively; adjusted OR 1.52 [0.84, 2.93] and 2.06 [0.91, 5.28] respectively), with a larger effect estimate in the latter case. Still, this association did not reach statistical significance in our analysis.

**Fig. 4.**
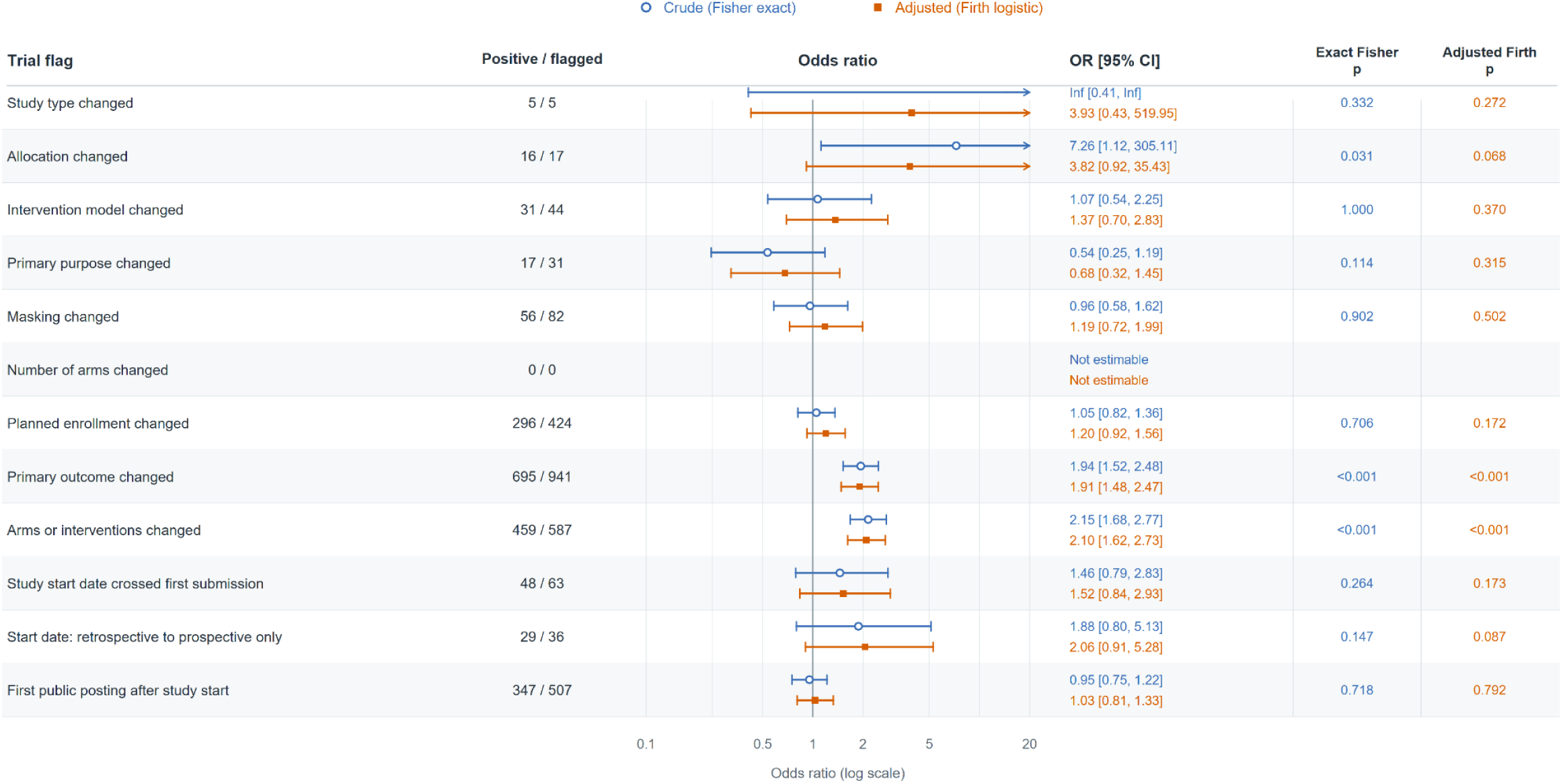
Association between primary endpoint positivity and flag categories. Blue open circles and orange squares represent crude and adjusted odds ratios (ORs), respectively; horizontal lines indicate 95% confidence intervals. Each flag was assessed separately, comparing trials with and without that flag. Crude and adjusted p-values derive from two-sided Fisher’s exact tests and penalized likelihood-ratio tests, respectively.

### Nested field change subtypes

We also conducted this analysis for the predefined subtypes of the flags “primary outcome changed” and “arms and interventions changed” (Fig. 5). The strongest association was estimated for the subtypes “primary outcome added” and “primary outcome removed” (unadjusted OR 1.83 [1.44-2.32] and 1.82 [1.44-2.31] respectively; adjusted OR 1.66 [1.30-2.12] and 1.66 [1.30, 2.12]), as well as for “arm added” and “arm removed” (unadjusted OR 2.40 [1.79-3.26] and 2.15 [1.61-2.91] respectively; adjusted OR 2.08 [1.55-2.81] and 1.84 [1.37-2.49]).

**Fig. 5.**
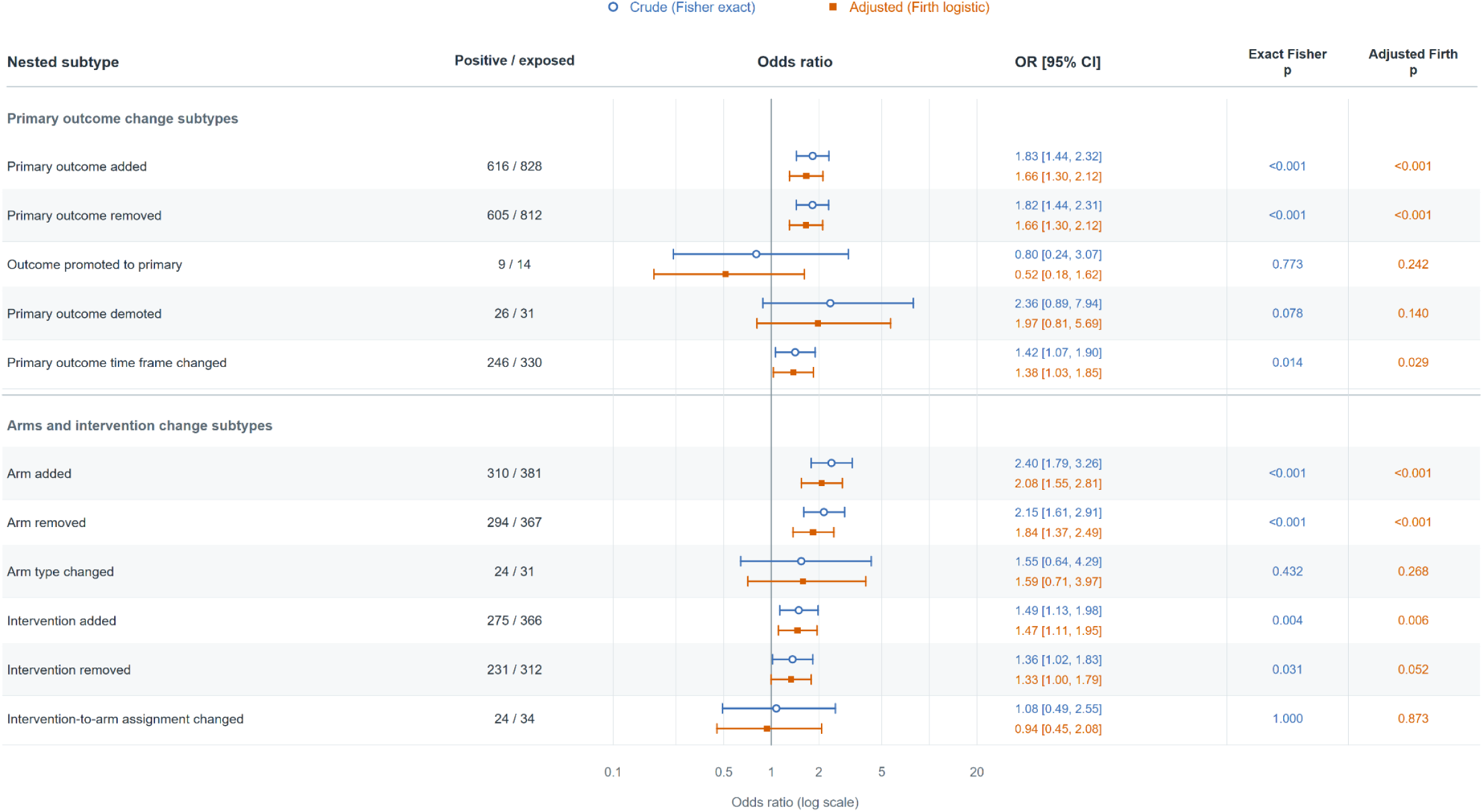
Association between primary endpoint positivity and nested flag subtypes.

### Endpoint positivity associations by timing relative to primary completion

The estimated unadjusted and adjusted ORs (Fig. 6) showed a markedly attenuated association in the “early only” group (changes made exclusively at or before the primary completion date) for both primary outcome changes and arms and interventions changes, with the effect estimate no longer reaching statistical significance in the former group.

**Fig. 6.**
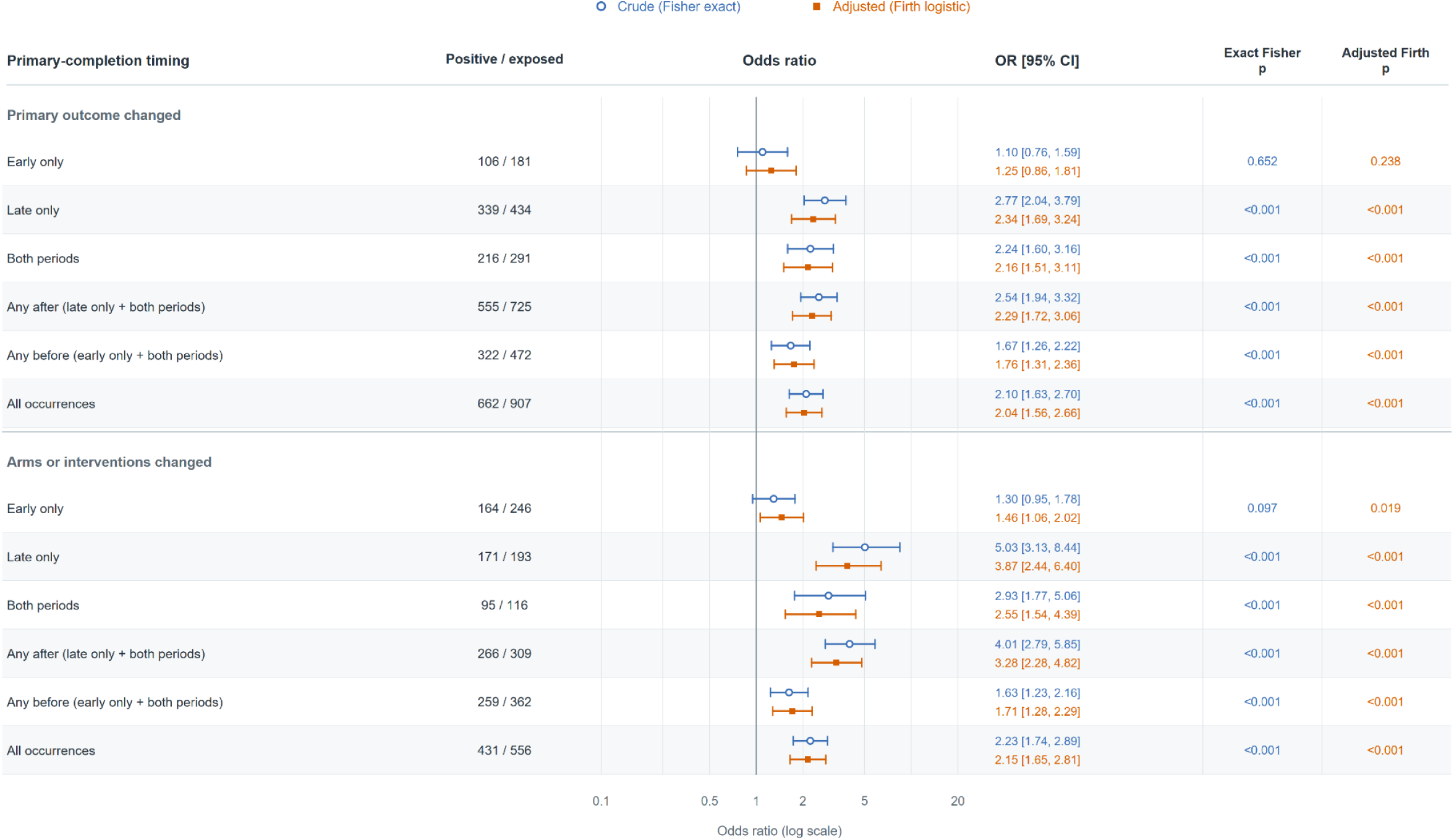
Association between primary endpoint positivity and nested flag categories by timing relative to primary completion date.

### LLM-flagged primary outcome changes

Our LLM-assisted semantic module identified 780 trials (Supplementary Table 3) with meaningful changes in the primary outcome (44.4% of the total). Compared to the deterministic pipeline a reduced frequency of outcome addition and removal was noted; many detected outcome changes were found distributed among new subcategories such as “definition or component”, “measurement method”, and “threshold”. Predictably, slightly more changes in outcome priority (promotions/demotions) could be detected by the LLM, since the deterministic pipeline required exact textual identity to establish a match.

The association with primary endpoint positivity which we observed for outcome changes identified by the deterministic pipeline was not evident in the case of changes classified using the LLM module. In the latter case unadjusted OR was 1.06 (0.84–1.35), and adjusted OR 1.15 (0.90–1.47).

When conducting the analysis by subcategories (see Supplementary figure 1), the only statistically significant association was with “outcome addition”, which resulted in an unadjusted OR of 1.89 (1.14-3.26) and an adjusted OR of 1.81 (1.10-3.11).

## Discussion

Our LLM-based abstract classification pipeline could effectively classify publications according to whether they reported RCT results and whether their primary endpoint was positive, as shown by our validation sample. Here perfect agreement with both human annotators was evident for RCT classification and perfect agreement was present between the LLM and one of the human annotators for endpoint positivity assessment, a result which confirms previously reported findings^16^.

Our automated assessment of registry changes in 1758 trials registered on ClinicalTrials.gov showed a particularly high prevalence of changes in the two nested fields we analyzed, primary outcomes and arms and interventions (69.3% and 44.7% respectively). This high result is attributable in part to the fact that we assessed for each trial every single version transition recorded on ClinicalTrials.gov, compared to most other comparable studies which only measured changes between two versions, or a limited number of versions at specific time landmarks. Another reason for the high prevalence of changes is the high sensitivity to textual changes of the deterministic pipeline we employed, which inevitably captures many minor text modifications such as orthographic corrections, alternative spelling or clarifications.

Outcomes between different versions can also only be matched if outcome type and measure are identical after text normalization, which probably leads to a high number of outcome modifications being classified as a pair of one addition + one removal.

We addressed this issue by developing an additional LLM-assisted semantic module for analysis of primary outcome changes. Similar approaches have already been tested in other works^17–19^ and show good discrimination capacity. The LLM-based approach by Ying et al.^19^ in particular showed remarkable accuracy in identifying additions, removals and changes of outcome priority, as well as in classifying changes of outcome definition. Our LLM-module found meaningful primary outcome changes in 44.4% of all trials. The prevalence of outcome additions, removals, and replacements (the latter representing the combination of existing outcome removal and new outcome addition) was 8.9%, 2.0%, and 4.3%, respectively. When changes to outcome time frames were also considered, the overall proportion of trials exhibiting at least one of these modifications reached 31.8%, which is remarkably similar to the prevalence of 30.8% found by Bulovic et al.^18^ when they assessed the sum of outcome addition, removal and time frame modification.

Our analysis also included two timing-related flags, “first post after study start” and “start date crossed first submission boundary”. “First post after study start” was flagged in 684 trials (38.9%) out of a total of 1758. All but three of these trials were submitted for registration after 2005, the year of the ICMJE deadline for prospective registration. This flag is thus a proxy for retrospective registration, although it considers the first public posting date and not the first submission date, which usually precedes it by a few days. A conservative estimate which applies a “grace period” of thirty days between the reported actual study start and the date of first public posting still results in 96 flagged trials (5.46% of the total). It is nevertheless possible that those trials were first registered in another ICMJE-accepted registry.

The actual study start date is not fixed and can be retrospectively revised on ClinicalTrials.gov, although all modifications are preserved in the registry history. The flag “start date crossed first submission boundary” identifies cases where the start date is moved beyond the boundary of the first submission date. Modification of start date from before to after the first submission date was observed in 80 trials (4.55% of all trials). Of these, 34 trials had a bidirectional crossing, which is compatible with a possible clerical error, and 46 trials (2.61%) had a crossing in the retrospective-to-prospective direction only. This is equivalent to a “retroactively prospective registration”, as defined by Holst et al.^20^, and is recognized as a potentially problematic practice. The prevalence of this phenomenon was 2.0% in the sample of 11,908 trials assessed by Holst, which is very similar to our result.

Primary outcome changes were positively associated with trial endpoint positivity in our analysis, a finding that is compatible with results from previous studies ^6,21^: Chen et al. reported an association between discrepancies in registered versus published primary outcomes and intervention effect size, whereas Florez et al. examined primary outcome changes across various sources (including ClinicalTrials.gov records and successive versions of study protocols) and identified an association with primary endpoint positivity, using an approach more closely resembling that of our study.

A new finding which, to the best of our knowledge, has not yet been reported in the literature is the positive association between changes in arms and interventions and trial success. Modifications to this registry section are still relatively understudied, with very few published reports^22^ on the frequency of such changes in the current version of ClinicalTrials.gov (where arms and interventions have been combined in one structured item).

The timing of changes to registry records is relevant, since changes made before enrollment of the first patient can represent harmless protocol refinements, whereas changes after study start warrant closer scrutiny. Changes made after primary completion date are particularly concerning, since they may potentially involve post hoc modifications to a prespecified study design. For this reason, we examined whether the strength of the associations between nested field changes and endpoint positivity varied according to their timing relative to primary completion. The associations were substantially weaker for trials with changes occurring only before primary completion but were most pronounced when trials had those changes occurring exclusively afterwards. This reinforces the hypothesis that modifications to study design late during a trial may introduce bias into results reporting. Ramagopalan et al. showed a similar finding, identifying an association between primary outcome changes occurring after primary completion and statistically significant primary outcomes ^23^, although they did not compare this association with primary outcome changes occurring before primary completion.

We also evaluated the association between the results of the primary outcome analyses conducted by our LLM-assisted module and endpoint positivity. In this case the association was unexpectedly weak and not statistically significant (unadjusted OR 1.06, adjusted OR 1.15). This might be partly because the LLM-module also takes into account the outcome description data element, which is ignored by the deterministic pipeline. This could make this module more sensitive to changes to outcome components such as measurement method or cutoff which, while semantically meaningful, are less methodologically salient than the type of changes considered “major” according to a definition first proposed by Chan et al.^24^, which only includes primary outcome additions, omissions and changes to outcome priority. This is reinforced by our finding in the subcategory analysis of a statistically significant association with LLM-detected primary outcome additions.

The present study contributes to the existing literature in several important ways: first, we conducted a systematic analysis of the complete registry histories in a large cohort of trials. Our information extraction extended beyond primary outcomes to include multiple textual fields describing study design characteristics. No previous study appears to have examined longitudinal changes across such a broad range of registry data elements and to have included all transitions between successive record versions. Second, we identified an association between the occurrence of changes to arms and interventions and trial positivity, a result which to our knowledge has not been reported before and extends previous findings which focused on selective reporting around primary outcome modifications. Third, we provided the first comparison of registry record changes according to their timing, revealing that changes after primary completion are more strongly associated with endpoint positivity. Our findings were also suggestive of a possible association with start date modifications that shift the apparent registration status from retrospective to prospective. These changes were previously characterized^20^, but have not been assessed yet in relation to trial positivity. Fourth, our pipeline provides a scalable framework which facilitates the automated extraction, parsing and analysis of registry data across large cohorts of trials for which manual assessment would otherwise be prohibitively time- and resource-intensive. This could serve as a useful tool for researchers investigating changes to trial registry records.

Our findings should be interpreted in light of several limitations: first, we restricted our analysis to a cohort of trials published in four major high-impact medical journals in a recent time interval. This could limit the generalizability of our results to other trials published in different journals, or in an earlier era. Second, our deterministic pipeline cannot fully distinguish semantically meaningful changes from minor textual modifications. We partly addressed this issue by having the flagging rules ignore textual modifications to the “description” data element, but we cannot exclude that by doing so we incurred in loss of information, or that we were still not fully able to filter out inconsequential changes. Third, we did not perform validation of our

LLM-assisted module for detecting outcome changes against a manually annotated sample. Previous research has demonstrated the high accuracy of generalist proprietary LLMs in performing this task^19^. However, we did not fully replicate their method because of cost-related feasibility constraints. Future work could be directed to refine this component of our pipeline by performing internal and external validation. Fourth, we only assessed changes within trial registry histories but did not examine discrepancies between registry records and corresponding publications, due to the complexity of using an automated workflow to extract this information from unstructured text. It is conceivable that further inconsistencies could emerge by extending the analysis in this manner.

Future research could extend our automated approach to other ClinicalTrials.gov fields and sections not included by our analysis: for example, eligibility criteria would be a candidate for further study. In a previous investigation, Cihoric et al. found that the readability and complexity of eligibility criteria were among the most informative predictors of trial withdrawal or termination in a model based on ClinicalTrials.gov data^25^. Future work could examine whether changes to these criteria over the course of a trial are associated with reported primary endpoint positivity. Overall, textual assessment of complex nested fields could be greatly facilitated by an LLM-assisted workflow. The approach we proposed for primary outcome changes could be further improved and also applied to the arms and interventions section. We conducted our study only on the ClinicalTrials.gov registry: an analogous evaluation could be performed for other registries belonging to the WHO International Clinical Trials Registry Platform (ICTRP). Finally, larger trial cohorts could allow more precise estimation of associations with endpoint positivity for additional variables for which our analysis was limited by the small size of the corresponding subgroups.

## Conclusions

This study revealed an association between primary endpoint positivity and changes to the primary outcomes and arms and interventions sections of trial registry records in a large cohort of RCTs, with the associations being most apparent when registry modifications occurred after primary completion date. These findings underscore the importance of transparent reporting of outcomes and other trial characteristics. The automated workflow we have developed for this analysis could facilitate future assessments of registry modifications at scale.

## Data Availability

All data and code used to obtain this study's results are available online at

https://github.com/purevdorj-ayurzana/rct_abstract_classifier

https://github.com/purevdorj-ayurzana/clinicaltrials_registry_pipeline

## Declarations

Ethics approval and consent to participate: Not applicable.

## Competing interests

The authors declare no conflict of interest.

## Funding/Support

This work was funded by the Swiss Cancer Research grant KFS-6477-08-2025

## Data and code availability statement

All data and code used to obtain this study’s results are available at https://github.com/purevdorj-ayurzana/rct_abstract_classifier and https://github.com/purevdorj-ayurzana/clinicaltrials_registry_pipeline

## Author contributions

Conceptualization: P.W.; Methodology: A.P., P.W.; Data curation: A.P., P.W.; Formal analysis: A.P.; Writing - original draft: A.P.; Writing - Review and editing. P.W., D.R.Z., F.D.; Supervision: P.W.; Project administration: P.W.

All authors read and approved the final manuscript.

## Notes

### Competing Interest Statement

The authors have declared no competing interest.

## References

1. Home | ClinicalTrials.gov [Internet]. [cited 2026 Nov 10]. Available from: https://clinicaltrials.gov/

2. WMA - The World Medical Association-WMA Declaration of Helsinki – Ethical Principles for Medical Research Involving Human Participants [Internet]. [cited 2026 Oct 5]. Available from: https://www.wma.net/policies-post/wma-declaration-of-helsinki/

3. FDAAA 801 and the Final Rule | ClinicalTrials.gov [Internet]. [cited 2026 Oct 5]. Available from: https://clinicaltrials.gov/policy/fdaaa-801-final-rule

4. Laine C, Horton R, DeAngelis CD, Drazen JM, Frizelle FA, Godlee F, et al. Clinical trial registration. BMJ. 2007 Jun 9;334(7605):1177–8. doi:10.1136/bmj.39233.510810.80 PubMed PMID: 17548363; PubMed Central PMCID: PMC1889969.

5. Huang Z, Zeng L, Ruan Z, Zeng Q, Yan H, Jiang W, et al. RETRACTED ARTICLE: Time-of-day immunochemotherapy in non-small cell lung cancer: a randomized phase 3 trial. Nat Med. 2026 Apr;32(4):1233–40. doi:10.1038/s41591-025-04181-w

6. Chen T, Li C, Qin R, Wang Y, Yu D, Dodd J, et al. Comparison of Clinical Trial Changes in Primary Outcome and Reported Intervention Effect Size Between Trial Registration and Publication. JAMA Netw Open. 2019 Jul 19;2(7):e197242. doi:10.1001/jamanetworkopen.2019.7242

7. Huić M, Marušić M, Marušić A. Completeness and Changes in Registered Data and Reporting Bias of Randomized Controlled Trials in ICMJE Journals after Trial Registration Policy. PLOS ONE. 2011 Sep 21;6(9):e25258. doi:10.1371/journal.pone.0025258

8. Mathieu S, Boutron I, Moher D, Altman DG, Ravaud P. Comparison of Registered and Published Primary Outcomes in Randomized Controlled Trials. JAMA. 2009 Sep 2;302(9):977–84. doi:10.1001/jama.2009.1242

9. Walker KF, Stevenson G, Thornton JG. Discrepancies between registration and publication of randomised controlled trials: an observational study. JRSM Open. 2014 Apr 9;5(5):2042533313517688. doi:10.1177/2042533313517688 PubMed PMID: 25057391; PubMed Central PMCID: PMC4012655.

10. Holst M, Haslberger M, Yerunkar S, Strech D, Hemkens LG, Carlisle BG. Frequency of multiple changes to prespecified primary outcomes of clinical trials completed between 2009 and 2017 in German university medical centers: A meta-research study. PLOS Med. 2023 Oct 31;20(10):e1004306. doi:10.1371/journal.pmed.1004306 PubMed PMID: 37906614; PubMed Central PMCID: PMC10645365.

11. Jones CW, Keil LG, Holland WC, Caughey MC, Platts-Mills TF. Comparison of registered and published outcomes in randomized controlled trials: a systematic review. BMC Med. 2015 Nov 18;13:282. doi:10.1186/s12916-015-0520-3 PubMed PMID: 26581191; PubMed Central PMCID: PMC4650202.

12. Ramagopalan S, Skingsley AP, Handunnetthi L, Klingel M, Magnus D, Pakpoor J, et al. Prevalence of primary outcome changes in clinical trials registered on ClinicalTrials.gov: a cross-sectional study. F1000Research. 2014 Mar 26;3:77. doi:10.12688/f1000research.3784.1 PubMed PMID: 25075294;

13. PubMed Central PMCID: PMC4032105.

13. Lamberink HJ, Vinkers CH, Lancee M, Damen JAA, Bouter LM, Otte WM, et al. Clinical Trial Registration Patterns and Changes in Primary Outcomes of Randomized Clinical Trials From 2002 to 2017. JAMA Intern Med. 2022 Jul 1;182(7):779–82. doi:10.1001/jamainternmed.2022.1551 PubMed PMID: 35575802; PubMed Central PMCID: PMC9112139.

14. Damen JA, Heus P, Lamberink HJ, Tijdink JK, Bouter L, Glasziou P, et al. Indicators of questionable research practices were identified in 163,129 randomized controlled trials. J Clin Epidemiol. 2023 Feb 1;154:23–32. doi:10.1016/j.jclinepi.2022.11.020

15. Carlisle BG. Analysis of clinical trial registry entry histories using the novel R package cthist. PLOS ONE. 2022 Jul 1;17(7):e0270909. doi:10.1371/journal.pone.0270909

16. Windisch P, Dennstädt F, Koechli C, Förster R, Schröder C, Aebersold DM, et al. A Pipeline for the Automatic Identification of Randomized Controlled Oncology Trials and Assignment of Tumor Entities Using Natural Language Processing [Internet]. medRxiv; 2024 [cited 2026 Oct 14]. p. 2024.07.01.24309767. Available from: https://www.medrxiv.org/content/10.1101/2024.07.01.24309767v1 doi:10.1101/2024.07.01.24309767

18. Ying X, Vorland CJ, Qureshi R, Brown AW, Kilicoglu H, Saldanha I, et al. GPT’s Performance in Identifying Outcome Changes on ClinicalTrials.gov [Internet]. MetaArXiv; 2024 [cited 2026 Oct 2]. Available from: https://osf.io/preprints/metaarxiv/npvwr_v1/ doi:10.31222/osf.io/npvwr

18. Bulovic I, Wunnava S, Yoon W, Dunn AG, Miller T, Bourgeois FT. Monitoring Changes in Clinical Trial Primary Outcomes Using Large Language Models. JAMA Netw Open. 2026 Sep 16;9(9):e2634214. doi:10.1001/jamanetworkopen.2026.34214

19. Ying X, Ninan K, Oberste JP, Vorland CJ, Li T, Brown AW, et al. An automated approach to improve clinical trial registration and to identify outcome changes on ClinicalTrials.gov. Npj Digit Med. 2026 Jul 29. doi:10.1038/s41746-026-03045-y

20. Holst M, Carlisle BG. Trials that turn from retrospectively registered to prospectively registered: a cohort study of “retroactively prospective” clinical trial registration using history data. Trials. 2024 Mar 14;25(1):189. doi:10.1186/s13063-024-08029-5 PubMed PMID: 38486299; PubMed Central PMCID: PMC10938677.

21. Florez MA, Jaoude JA, Patel RR, Kouzy R, Lin TA, De B, et al. Incidence of Primary End Point Changes Among Active Cancer Phase 3 Randomized Clinical Trials. JAMA Netw Open. 2023 May 1;6(5):e2313819. doi:10.1001/jamanetworkopen.2023.13819 PubMed PMID: 37195664; PubMed Central PMCID: PMC10193180.

22. Gujinović D, Viđak T, Melnikova M, de Sousa NJF, Marušić A. Changes in Registration Parameters for Ongoing Clinical Trials in Ukraine After 2022 Russian Invasion. JAMA Netw Open. 2023 Jun 26;6(6):e2320202. doi:10.1001/jamanetworkopen.2023.20202

23. Ramagopalan SV, Skingsley AP, Handunnetthi L, Magnus D, Klingel M, Pakpoor J, et al. Funding source and primary outcome changes in clinical trials registered on ClinicalTrials.gov are associated with the reporting of a statistically significant primary outcome: a cross-sectional study. F1000Research. 2015 Apr 24;4:80. doi:10.12688/f1000research.6312.2 PubMed PMID: 26069729; PubMed Central PMCID: PMC4431380.

24. Chan AW, Krleza-Jerić K, Schmid I, Altman DG. Outcome reporting bias in randomized trials funded by the Canadian Institutes of Health Research. CMAJ Can Med Assoc J J Assoc Medicale Can. 2004 Sep 28;171(7):735–40. doi:10.1503/cmaj.1041086 PubMed PMID: 15451835; PubMed Central PMCID: PMC517858.

25. Cihoric N, Gavric S, Dennstädt F, Jovanovic A. A multi-factor machine learning model for predicting and characterizing clinical trial failures. Discov Artif Intell. 2026 Aug 15;6(1):804. doi:10.1007/s44163-026-01965-y

