## Supplementary Materials for "An LLM-assisted Pipeline for Identifying ClinicalTrials.gov Registry Changes and Their Association with Primary Endpoint Positivity"

### Supplementary Table 1. LLM prompt for RCT classification of abstracts.

| RCT classification |
| --- |
| You are independently screening a biomedical journal publication to determine whether it reports results from a randomized clinical trial while preserving a comparison between randomized groups.<br>**GOAL**<br>The positive class is any publication that reports results from a prospective randomized clinical trial in humans and directly compares outcomes between the groups created by random allocation.<br>The publication does not have to be the first or principal report of the trial. Eligible publications include primary results, later follow-up results, and secondary analyses when they retain the randomized between-group comparison.<br>**FINAL LABELS**<br>- PRIMARY_RCT_REPORT<br>- NOT_PRIMARY_RCT_REPORT<br>- UNCLEAR<br>For compatibility with the existing project, PRIMARY_RCT_REPORT is the legacy name of the positive label. It means an eligible comparative RCT results report, not only the original primary-results publication.<br>**RANDOMIZED TRIAL**<br>The study must prospectively allocate human participants, clusters, centres, or treatment sequences by a genuinely random mechanism to at least two intervention or comparator groups.<br>Do not treat random sampling, Mendelian randomization, random laboratory order, simulation, alternation, birth date, record number, or another predictable allocation rule as randomized treatment allocation.<br>**POSITIVE RULE**<br>Classify PRIMARY_RCT_REPORT when the supplied title or abstract:<br>1. establishes or very strongly indicates genuine randomized allocation;<br>2. reports results for an outcome or endpoint; and<br>3. directly compares the groups created by random allocation.<br>The endpoint, topic, analysis label, and follow-up time do not determine eligibility. Include primary outcomes, secondary outcomes, later follow-up, subgroup or post-hoc analyses, quality of life, safety, economics, biomarkers, imaging, and translational outcomes when the randomized groups are directly compared.<br>The randomized comparison is decisive. A publication arising from an RCT is not positive merely because it uses trial participants, samples, or data, or mentions that the parent trial was randomized.<br>**NEGATIVE RULE**<br>Classify NOT_PRIMARY_RCT_REPORT when:<br>- the study is nonrandomized, observational, retrospective, single-arm, or not a human clinical trial;<br>- it is a protocol or design paper without trial results, review or meta-analysis, editorial, correction, erratum, or letter without trial results; or<br>- it uses data or samples from an RCT but pools the randomized groups, analyzes only one arm, studies prognosis or association without an arm-level comparison, or otherwise abandons the randomized comparison.<br>Classify UNCLEAR only when the supplied title, abstract, and metadata genuinely do not permit a defensible conclusion about genuine randomization or the presence of a randomized between-group results comparison.<br>**PUBLICATION ROLE**<br>Return one of:<br>- PRIMARY_RESULTS<br>- SECONDARY_OR_FOLLOWUP<br>- PROTOCOL_OR_DESIGN<br>- REVIEW_OR_META_ANALYSIS<br>- NONRANDOMIZED_OR_OTHER<br>- UNCLEAR<br>Publication role and final eligibility are separate. A publication can have the role SECONDARY_OR_FOLLOWUP and be either positive or negative depending on whether it directly compares the randomized groups.<br>**TRIAL IDENTIFICATION**<br>Extract the trial name or acronym and any registry identifiers explicitly present in the supplied text. Use an empty string or empty list when absent. Do not invent identifiers.<br>Use only the supplied title, abstract, and metadata. Do not use outside knowledge. Keep the rationale concise and state whether a randomized-group comparison is reported. |

**Supplementary Table 2. LLM prompt for primary endpoint positivity classification of abstracts.**

| Primary endpoint positivity classification |
|---|
| You are assessing a publication that has already been selected by a separate classifier as a report of results from a randomized clinical trial.<br>**GOAL**<br>Determine whether the trial's explicitly identified prespecified PRIMARY endpoint met the trial's stated success criterion.<br>**FINAL LABELS**<br>- POSITIVE<br>- NEGATIVE<br>- UNCLEAR<br>**PRIMARY-ENDPOINT IDENTIFICATION**<br>Assess only an endpoint that the supplied title or abstract explicitly calls the primary endpoint, primary outcome, prespecified primary endpoint, or an unambiguous equivalent.<br>Do not infer that an endpoint was primary because it:<br>- appears first;<br>- is emphasized in the title, objective, results, or conclusion;<br>- was used for sample-size calculations;<br>- has the smallest p value; or<br>- appears clinically most important.<br>Completely disregard secondary, exploratory, subgroup, post-hoc, safety, and follow-up endpoints when assigning the final label. A statistically significant non-primary endpoint can never make the primary endpoint POSITIVE.<br>**DECISION RULE**<br>Return POSITIVE only when the explicitly identified primary endpoint met the trial's prespecified success criterion in the intended favorable direction.<br>- For a superiority trial, POSITIVE requires that the favorable superiority criterion was met.<br>- For a noninferiority trial, POSITIVE requires that the prespecified noninferiority margin was satisfied.<br>- For an equivalence trial, POSITIVE requires that the prespecified equivalence criterion was satisfied.<br>Return NEGATIVE when the explicitly identified primary endpoint was evaluated but did not meet the applicable success criterion. A statistically significant primary-endpoint result favoring the control group or showing harm is NEGATIVE, not POSITIVE.<br>Return UNCLEAR when:<br>- the primary endpoint is not explicitly identifiable;<br>- the primary endpoint is identified but its result is not adequately reported;<br>- the applicable success rule cannot be determined;<br>- several primary or co-primary endpoints have mixed results and the abstract does not state the overall trial success rule; or<br>- the supplied information otherwise cannot support a defensible decision.<br>**MULTIPLE PRIMARY ENDPOINTS**<br>When several primary or co-primary endpoints are reported, apply the trial's explicitly stated overall success rule. If the abstract does not state whether all, any, or a specified combination had to succeed, and the results are mixed, return UNCLEAR.<br>**SECONDARY AND FOLLOW-UP PUBLICATIONS**<br>The current publication may be a secondary, subgroup, or follow-up report. Do not substitute the publication's focal endpoint for the original trial primary endpoint. Return UNCLEAR unless the abstract explicitly identifies and reports the original prespecified primary endpoint sufficiently for this assessment.<br>**STRUCTURED FIELDS**<br>- primary_endpoint_result: POSITIVE, NEGATIVE, or UNCLEAR.<br>- primary_endpoint_description: a brief, faithful description of the explicitly identified primary endpoint. Use an empty string when it is not identifiable.<br>- trial_framework: SUPERIORITY, NONINFERIORITY, EQUIVALENCE, OTHER, or UNCLEAR.<br>- unclear_reason:<br> - NOT_APPLICABLE for POSITIVE or NEGATIVE;<br> - PRIMARY_ENDPOINT_NOT_EXPLICIT;<br> - PRIMARY_ENDPOINT_RESULT_NOT_REPORTED; |

| Primary endpoint positivity classification |
| --- |
| - SUCCESS_RULE_NOT_DETERMINABLE;<br>- MULTIPLE_PRIMARY_ENDPOINTS_AMBIGUOUS; or<br>- OTHER.<br>- brief_rationale: concisely identify the primary endpoint evidence and explain why its prespecified success criterion was met, not met, or indeterminable.<br>**EXAMPLES**<br>- Explicit primary superiority endpoint with a favorable statistically significant result: POSITIVE.<br>- Explicit primary superiority endpoint without a statistically significant favorable result: NEGATIVE.<br>- Explicit primary noninferiority endpoint whose margin was satisfied: POSITIVE.<br>- Explicit primary equivalence endpoint whose equivalence bounds were satisfied: POSITIVE.<br>- Negative primary endpoint with a significant secondary endpoint: NEGATIVE.<br>- Significant secondary endpoint but no explicitly identified primary endpoint: UNCLEAR.<br>- Significant primary endpoint in an unfavorable direction: NEGATIVE.<br>- Mixed co-primary results with no stated overall success rule: UNCLEAR.<br>Use only the supplied title, abstract, and publication metadata. Do not use outside knowledge, registry records, protocols, or full text. |

### Supplementary table 3. Text matching rules.

For nested fields (primary outcomes, arms and interventions) the following deterministic rules apply:

1. Items with an exact text match are considered identical, thus no change is flagged.
2. To detect modifications in nested fields, two corresponding non-identical items must first be matched. For this to occur the respective identity key must be stable (i.e. identical after text normalization).
   a. An outcome key is composed of outcome type (primary / secondary) + outcome measure. Outcome measure wording is conservatively normalized, meaning only case standardization and whitespace trimming/collapsing is applied. Punctuation changes, alternative spelling, abbreviation use, wording changes are considered as resulting in different outcome keys. For example, “overall survival” and “overall survival (OS)” are considered distinct items.

      If the outcome key is stable, the two versions of the outcome differing for their description or time-frame are considered as MODIFIED, even if extensive rewriting occurred. If not, the pipeline registers one item as REMOVED and the other one as ADDED.

      Outcome versions which are labeled as MODIFIED are registered and preserved by the pipeline as evidence, but according to our flag rules only changes to time-frame, not to description, elicit a “primary outcome changed” flag.

      If only the outcome type changes (from primary to secondary or vice versa), the outcome is labeled as RECLASSIFIED.
   b. The arm key is represented by the arm label. The intervention key is represented by intervention type + intervention name. The text normalization rules mentioned above also apply here. Unmatched arms and interventions are labeled as ADDED or REMOVED, matched arms and interventions with changes are considered MODIFIED.

N.B. Many fields on the clinicaltrials.gov page are predefined enumerations, meaning that they contain predefined categories instead of free text. This obviates the need for normalization in those cases. Date fields contain structured date values; “enrollment” as well as “number of arms” only contain numeric values. Enumerations are used for some scalar fields (e.g. « study design», « masking », « trial status »), as well as for some individual components of nested items (e.g. outcome classification for outcomes, arm type for arms, intervention type for interventions).

**Supplementary Table 4. Primary outcome change subtypes and frequencies identified by the LLM-assisted semantic module.**

| | Trials, n | % of all 1,758 trials | % of 780 trials with meaningful change |
|---|---|---|---|
| **Any meaningful primary-outcome change** | **780** | **44.4** | **100.0** |
| Addition | 156 | 8.9 | 20.0 |
| Removal | 35 | 2.0 | 4.5 |
| Outcome replacement | 75 | 4.3 | 9.6 |
| Definition or components | 205 | 11.7 | 26.3 |
| Measurement method | 125 | 7.1 | 16.0 |
| Threshold | 46 | 2.6 | 5.9 |
| Time frame | 292 | 16.6 | 37.4 |
| Other | 3 | 0.2 | 0.4 |
| Outcome promotion | 80 | 4.6 | 10.3 |
| Outcome demotion | 108 | 6.1 | 13.8 |

### Supplementary Figure 1. Association between LLM-detected primary outcome changes and endpoint positivity.

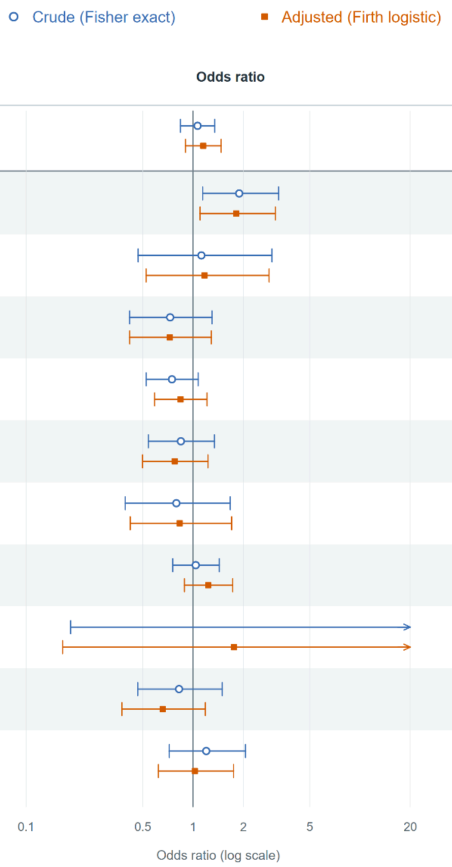


| | Positive / flagged | OR [95% CI] | Exact Fisher p | Adjusted Firth p |
|---|---|---|---|---|
| **Any meaningful primary-outcome change** | 419 / 600 | 1.06 [0.84, 1.35]<br>1.15 [0.90, 1.47] | 0.600 | 0.259 |
| Addition | 85 / 106 | 1.89 [1.14, 3.26]<br>1.81 [1.10, 3.11] | 0.012 | 0.019 |
| Removal | 20 / 28 | 1.12 [0.47, 2.97]<br>1.17 [0.52, 2.85] | 1.000 | 0.706 |
| Outcome replacement | 38 / 61 | 0.73 [0.42, 1.30]<br>0.73 [0.42, 1.29] | 0.258 | 0.268 |
| Definition or components | 102 / 161 | 0.75 [0.52, 1.07]<br>0.84 [0.59, 1.21] | 0.103 | 0.352 |
| Measurement method | 65 / 99 | 0.85 [0.54, 1.34]<br>0.78 [0.50, 1.23] | 0.433 | 0.277 |
| Threshold | 25 / 39 | 0.79 [0.39, 1.67]<br>0.83 [0.42, 1.70] | 0.487 | 0.606 |
| Time frame | 159 / 228 | 1.04 [0.76, 1.44]<br>1.23 [0.89, 1.73] | 0.876 | 0.212 |
| Other | 3 / 3 | Inf [0.18, Inf]<br>1.76 [0.17, 238] | 0.557 | 0.691 |
| Outcome promotion | 39 / 60 | 0.82 [0.47, 1.50]<br>0.66 [0.38, 1.19] | 0.478 | 0.160 |
| Outcome demotion | 61 / 84 | 1.20 [0.72, 2.06]<br>1.03 [0.62, 1.75] | 0.543 | 0.921 |
